# Return to work and care use by long-COVID patients on Bonaire, Caribbean Netherlands: Are patients their needs met?

**DOI:** 10.1101/2024.07.24.24309156

**Authors:** DSF Berry, G. Marchena, I. Tiemessen, A. Vegh, L. Jaspers, E. Geubbels

## Abstract

**Background:** Symptoms persisting ≥ 4 weeks after an acute SARS-CoV-2 infection (post-COVID condition, PCC) can have substantial consequences for the daily functioning and labor force participation of working age patients. We examined care consultations during PCC among patients on Bonaire, Caribbean Netherlands, identified activity limitations among patients who returned to work (RTW), and described the support that these patients indicated has or would have facilitated them in recovering and re-integrating.

**Methods:** Bivariate logistic regression was used to examine correlates of care consultations during PCC among 157 PCC patients on Bonaire, and to identify factors related to RTW among a subgroup of working age PCC patients with pre-pandemic employment (n=129). We applied in-depth qualitative analyses to assess patients’ lived experiences related to RTW and care during PCC.

**Results:** We found 37% of PCC patients consulted at least one (para)medical professional during PCC, of whom one in three consulted multiple professionals. 26% of patients recovered within one month since their acute infection; for patients unrecovered at time of interview, median time since infection was 249 days (IQR 30). Patients with comorbidity (OR=2.90, p<0.01, 95% CI [1.40-5.53]), under care prior to the pandemic (OR=2.52, p<0.01, 95% CI [1.30-4.95]), who had been hospitalized during the acute phase (OR=9.16, p<0.01, 95% CI [3.41, 29.21]), and patients who were aware that their insurance covered certain medical costs related to COVID-19 (after)care (OR=3.99, p<0.01, 95% CI [1.87, 8.81]) were more likely to consult care during PCC. Of patients who RTW (81%), 13% were working reduced hours, 35% experienced worsening of their symptoms after carrying out any physical or cognitive activity, and 40% encountered situations where they were unable to carry out work activities since they had COVID-19. Outside of the workplace, about one in four patients faced issues in sports activities and maintaining social relations since having COVID-19.

**Discussion:** Our findings highlight various factors related to care use during PCC by patients on Bonaire and patient identified needs related to support in the workplace and COVID-19 aftercare. Improving PCC related occupational and healthcare policies on Bonaire may prove beneficial for recovering patients their vitality and socio-economic vigor.

## BACKGROUND

By end of May 2020, roughly 18,755 SARS-CoV-2 cases and 631 related deaths were reported in the Caribbean [1]. The pandemic exacerbated various vulnerabilities in small island developing states (SIDS) in the region [2]. Concurrent to its population health impact, the response to the global pandemic disrupted regular healthcare services across many Latin American & Caribbean (LAC) nations, challenging the fragile health systems of SIDS with limited resources to respond to acute health emergencies, such as the Dutch Caribbean, to meet essential needs of the population [1,3–5]. Furthermore, the pandemic had a devastating impact on the economies of many islands in the LAC region [2–4,6]. Public health regulations installed to contain COVID-19 substantially affected sectors and industries that were critical for generating income and employment [3,7], and it is estimated that between February and April 2020, up to 700.000 jobs were lost in the Caribbean region [2].

While most individuals who have been infected with SARS-CoV-2 recover within a few weeks, persisting symptoms of COVID-19 (“Post COVID-19 Condition” (PCC)) can create substantial consequences for the daily functioning and labor force participation of working age patients [8]. Though most patients succeed at resuming work in their previous role, others are presented with challenges when faced with a relapsing pattern of symptoms or when existing symptoms worsen; having serious implications for job security, work productivity, and occupational safety and health [4,8,9].

Studies reporting on PCC related healthcare utilization and labor force participation in LAC settings, such as the Dutch Caribbean, remain scarce [5]. In May 2021, local physicians, employers, and policy makers on Bonaire, Dutch Caribbean, expressed experiencing trouble in identifying and accommodating the needs of these patients on their road to recovery, as much like elsewhere [10] the absence of evidence on effective treatments and programs prevented them from developing appropriate COVID-19 aftercare. Consequently, health services and insurance policies have not been optimally tailored to the needs of the patient population on Bonaire, which may have resulted in patients returning to work (RTW) prior to recovering where there was a substantial financial need or concern of job security [8].

This study aims to examine the trajectory of recovery of PCC patients on Bonaire by describing care consultations for persisting COVID-19 symptoms during PCC, identifying activity limitations among the subset of patients who RTW, and describing the healthcare provision and occupational support that has or would have facilitated these patients in recovering and re-integrating to work, using retrospective cohort data from Bonairean patients with disease onset between March 2020 and October 2021.

## METHODS

### Study design and data sources

We conducted a retrospective cohort study among symptomatic SARS-CoV-2 patients in Bonaire, Caribbean Netherlands [11]. To be eligible, participants had to reside in Bonaire, have access to a mobile phone, and be able to comprehend Papiamentu (local Creole), Dutch, English, or Spanish and be registered in the patient registry kept by the Bonairean municipal health service. Data was collected through telephone interviews between 15 November and 4 December 2021, after obtaining verbal informed consent. Respondents were asked to detail the presence and severity of 14 symptoms pre-infection, within the first four weeks after disease onset, and throughout the entire post-acute phase, plus a number of other questions about their medical and work history, their health seeking behavior, their functional status and their work reintegration trajectory. The full methodology of this study has been described elsewhere [11]. The Central Committee on Research Involving Human Subjects (CCMO Netherlands) confirmed on 8 October 2021 that the study did not require ethical approval due to its observational design.

### Study population

We included all laboratory confirmed SARS-CoV-2 positive cases who fit our case definition for PCC: “an individual with a laboratory confirmed SARS-CoV-2 positive test result, of whom at least one symptom self-attributed to the experienced SARS-CoV-2 infection lasted longer than four weeks” for whom data on care consultations during PCC was available (n=157). This is three patients fewer than the 160 PCC patients in the overarching study, because data on care consultations during PCC was not available for three PCC patients. Analyses focusing on RTW only included patients of the legal working age of <15 or >74 years on Bonaire (n=149) and with pre-pandemic employment (n=130), which was determined by work status at the time of interview. Of working age PCC patients with pre-pandemic employment (n=129), 14 cases who did not RTW and 11 patients for whom RTW status was missing (reasons unknown) were excluded from analyses characterizing RTW, so final RTW analyses included 104 PCC patients.

### Outcomes

Our main outcome variable included care consultations during PCC, defined as consulting at least one (para)medical professional for COVID-19 symptoms persisting at least four weeks after disease onset. For a full list of included professionals, see **Supplementary Table 1**. Patients were asked whether they had visited a (para)medical healthcare professional and if so, in what frequency. Our second outcome of interest concerned return to work status at time of interview (yes/no).

### Covariates

Demographic variables included gender, age, education level, household composition, and comorbidity; defined as presence of at least one diagnosed underlying condition. Comorbidities mentioned in open ended questions were recoded into the binary variable. Age was treated as a continuous variable. A patient’s pre-pandemic phase was defined as the situation before start of the COVID pandemic; the acute phase as the first four weeks after a SARS-CoV-2 infection and the post-acute phase as the period beyond those four weeks. For a full description of included covariates, see **Supplementary Table 1**.

### Statistical analysis

#### Descriptive analyses

We described pre-pandemic, acute, post-acute health care utilization, and return to work status. We presented categorical data using counts and frequencies and presented continuous data using mean (+SD) or median (IQR) where appropriate. We assessed the correlation between demographic- and health factors, and healthcare utilization using Fisher’s exact test or Wilcoxon test where appropriate (results not shown), and subsequently by using univariate logistic regression. All analyses were carried out in R version 4.1.3 (R Foundation for Statistical Computing).

#### Qualitative analyses

Open ended questions were analyzed to gain insight into traditional healing utilization, self-care practices, unmet health care needs and barriers, and into patients’ considerations around their decision to return to work when not feeling fully recovered. Two researchers performed thematic analyses of survey responses to open ended questions, of whom one, a public health nurse born and raised on Bonaire, was based on Bonaire and one, an epidemiologist with a background in nursing who was raised on the neighboring island Curaçao, was based in the Netherlands. Findings from the inductive approach based on Braun and Clarke’s methodology [12] were compared over two virtual meetings in November 2022. Final codes and themes and interpretation were determined during a third meeting with both researchers and the senior researcher early December 2022. All themes were included in the results.

## RESULTS

### Characteristics of PCC patients

Thus, a total of 157 patients (median age 43 years old) were included in this study (**Table 1**). A lower median age was observed among patients who did not consult care during PCC. The majority of patients were female (71%), employed (83%), living with their partner (24%) or partner and kids (24%), and had no underlying comorbidity (71%).

**Table 1.**
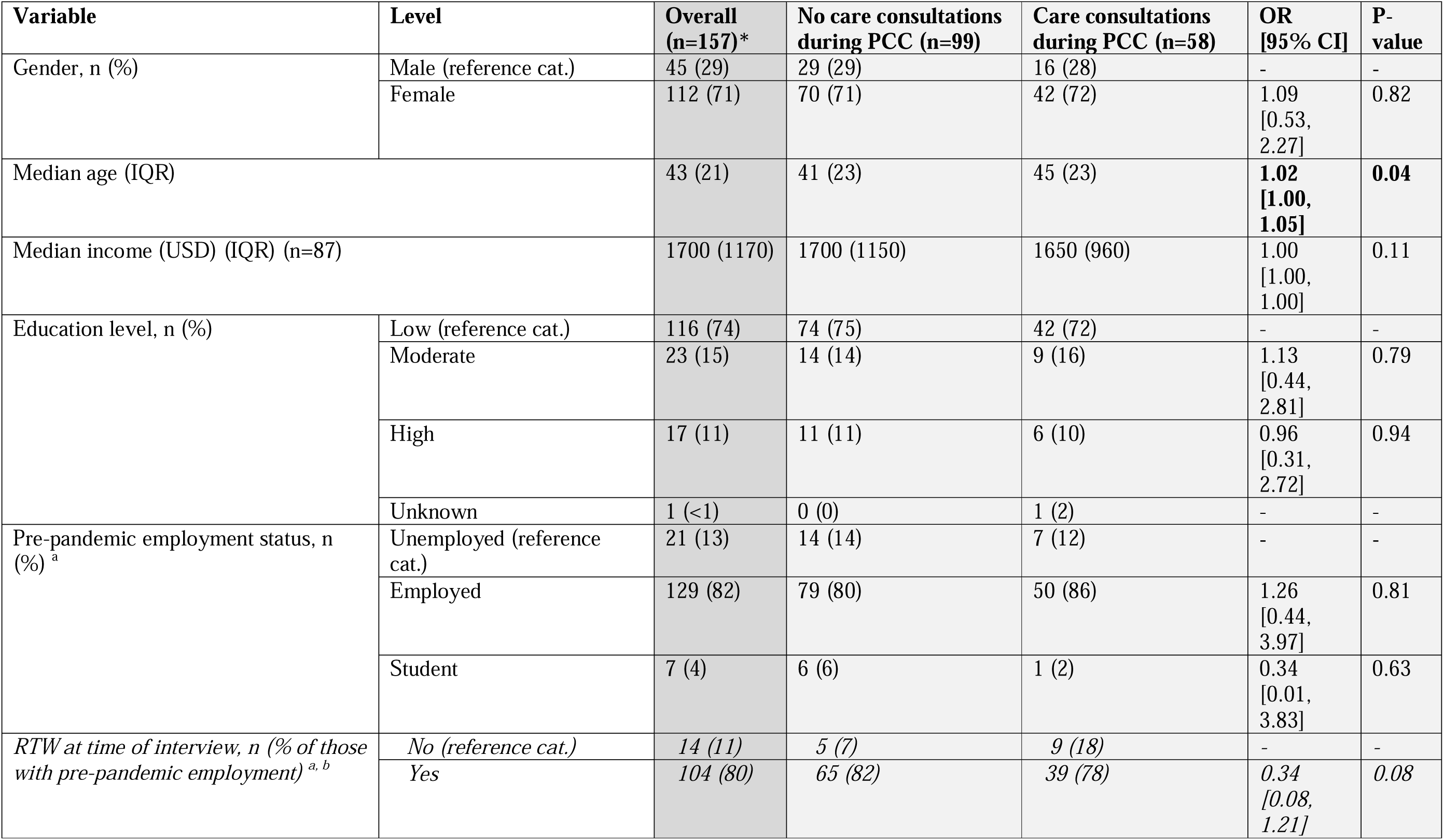
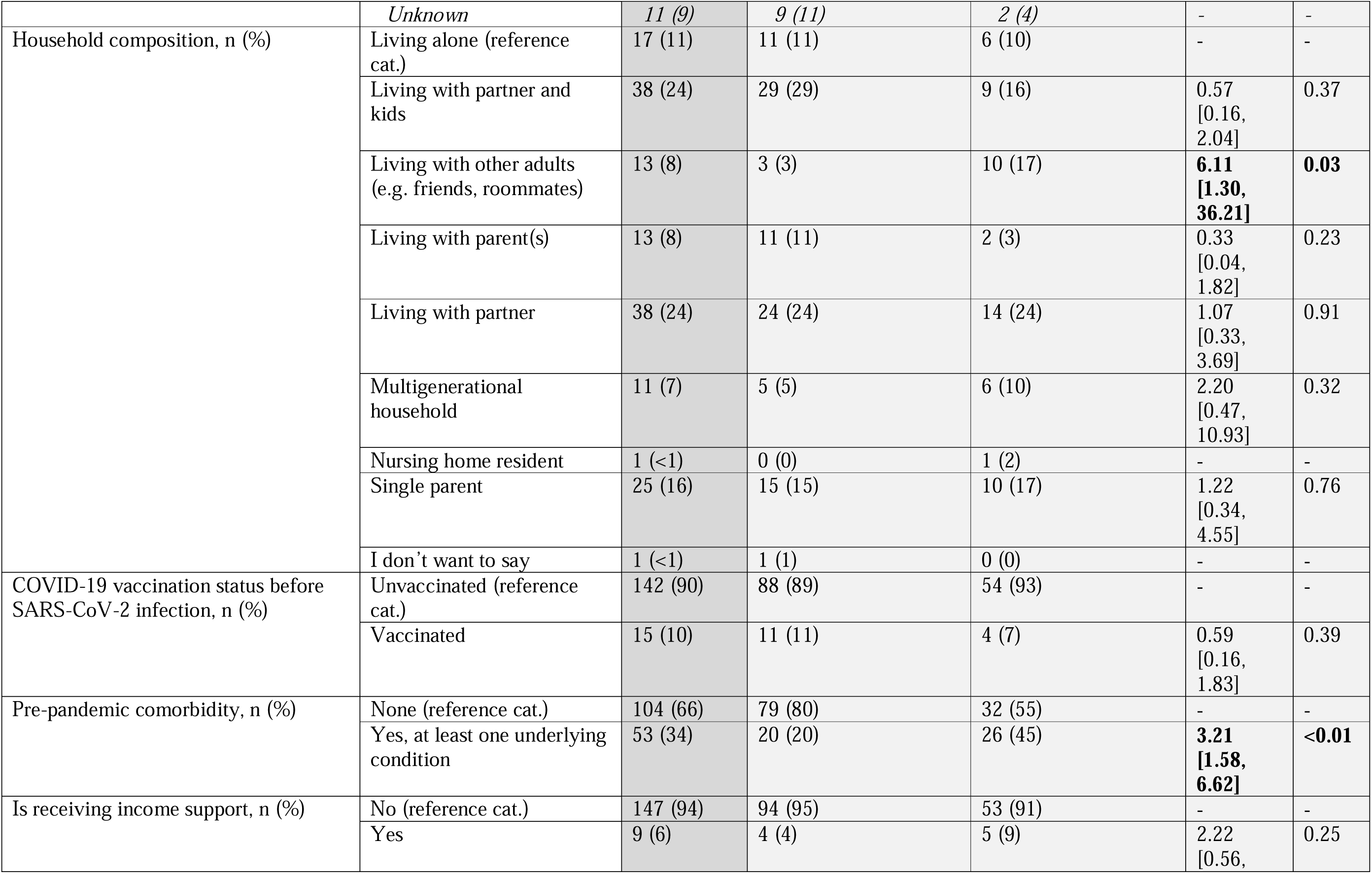
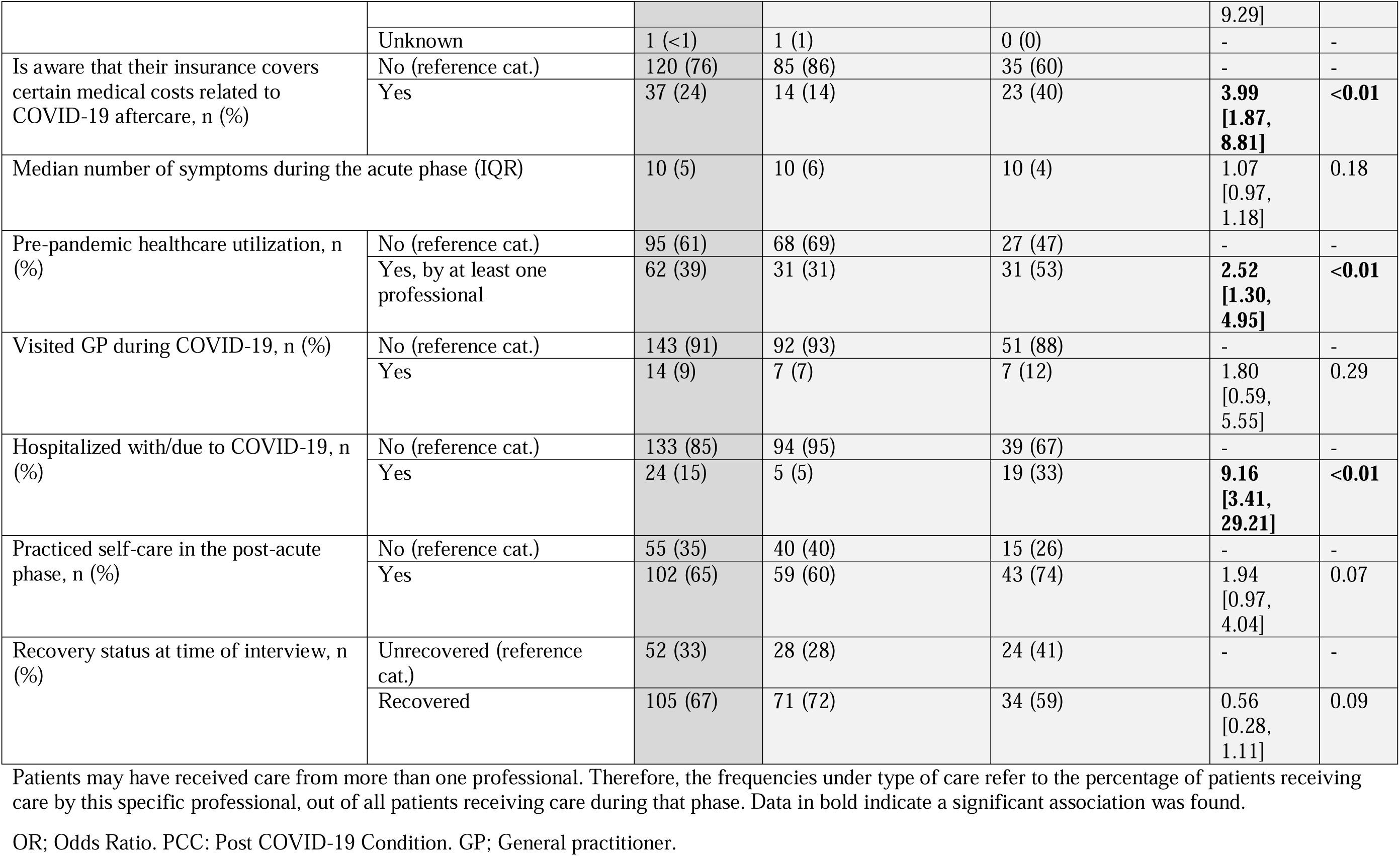
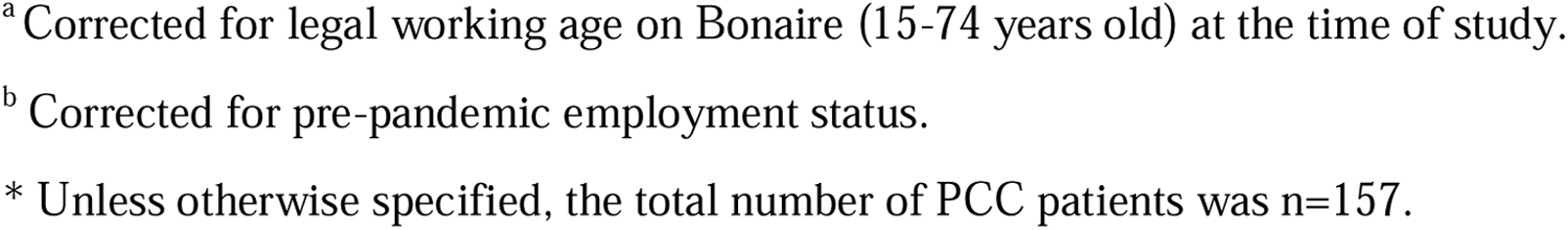
Baseline characteristics and factors associated with reported healthcare use beyond the acute phase by 157 PCC patients on Bonaire, Caribbean Netherlands, who tested positive for SARS-CoV-2 between 1 March 2020 and 1 October 2021.

### Healthcare utilization

#### Pre-pandemic & acute phase healthcare use

Prior to the pandemic, 39% of patients were receiving care by at least one professional (**Table 2**). Patients under care were most often visiting their GP (54%), psychologist (19%), or an internist (17%). During the acute phase, 9% of patients sought medical care from their GP and 15% were admitted to the hospital with or due to COVID-19 (**Table 1**). Common symptoms among patients visiting the GP in the initial four weeks included fatigue (93%), reduced physical endurance (93%), and sleeping problems (93%) (**Supplementary Table 2**). Among patients hospitalized with or due to COVID-19 who later developed PCC, common symptoms included reduced muscle strength (71%), reduced physical endurance (88%), and shortness of breath (79%).

**Table 2.** Care consultations prior to the pandemic and during post COVID-19 condition (PCC) for patients who tested positive for SARS-CoV-2 between 1 March 2020 and 1 October 2021, on Bonaire, Caribbean Netherlands, by type of healthcare provider (n=157).

|  | Prepandemic healthcare use, n (%) | Care consultations during PCC, n (%) |
| --- | --- | --- |
| No | 95 (61) | 99 (63) |
| Yes | 62 (39) | 58 (37) |
| *Cardiologist* | 6 (10) | 4 (7) |
| *Dermatologist* | 2 (3) | 0 (0) |
| *Diabetes nurse* | 1 (2) | 0 (0) |
| *ENT doctor* | 1 (2) | 1 (2) |
| *Gastroenterologist* | 2 (3) | 0 (0) |
| *GP* | 34 (54) | 39 (67) |
| *Gynecologist* | 6 (10) | 1 (2) |
| *HIV specialist* | 1 (2) | 0 (0) |
| *Home care professional* | 1 (2) | 6 (10) |
| *Immunologist* | 1 (2) | 0 (0) |
| *Insurance doctor* | 0 (0) | 2 (3) |
| *Internist* | 11 (17) | 6 (10) |
| *Manual therapist* | 2 (3) | 0 (0) |
| *Neurologist* | 2 (3) | 0 (0) |
| *Occupational doctor* | 2 (3) | 22 (38) |
| *Occupational therapist* | 1 (2) | 0 (0) |
| *Oncologist* | 1 (2) | 0 (0) |
| *Ophthalmologist* | 5 (8) | 0 (0) |
| *Orthopedist* | 1 (2) | 0 (0) |
| *Pediatrician* | 1 (2) | 0 (0) |
| *Physiotherapist* | 6 (10) | 15 (26) |
| *Podiatrist* | 1 (2) | 0 (0) |
| *Pulmonologist* | 2 (3) | 6 (10) |
| *Psychologist* | 12 (19) | 9 (16) |
| *Rheumatologist* | 2 (3) | 0 (0) |
| *Social worker* | 0 (0) | 1 (2) |
| *Specialist pain management* | 1 (2) | 0 (0) |
| *Surgeon* | 2 (3) | 0 (0) |
| *Urologist* | 2 (3) | 0 (0) |

Note: For a full list of definitions per variable, see **Supplementary Table 1**.
PCC; Post COVID-19 Condition. GP; General practitioner. ENT; Ear, nose and throat. HIV; Human Immunodeficiency Virus.

#### Care consultations during PCC

37% of patients consulted care during PCC. In total, 12 different professionals were consulted for persisting symptoms (**Figure 1**), most common being the GP (consulted by 67% of patients), occupational doctor (38%), and physiotherapist (26%) (**Table 2**). Specialist care, including the pulmonologist, internist, gynecologist, ENT doctor, or cardiologist, was consulted by 31% of patients. One in three patients consulting care during PCC had a history of hospital admission during the acute phase (**Table 1**).

**Figure 1.**
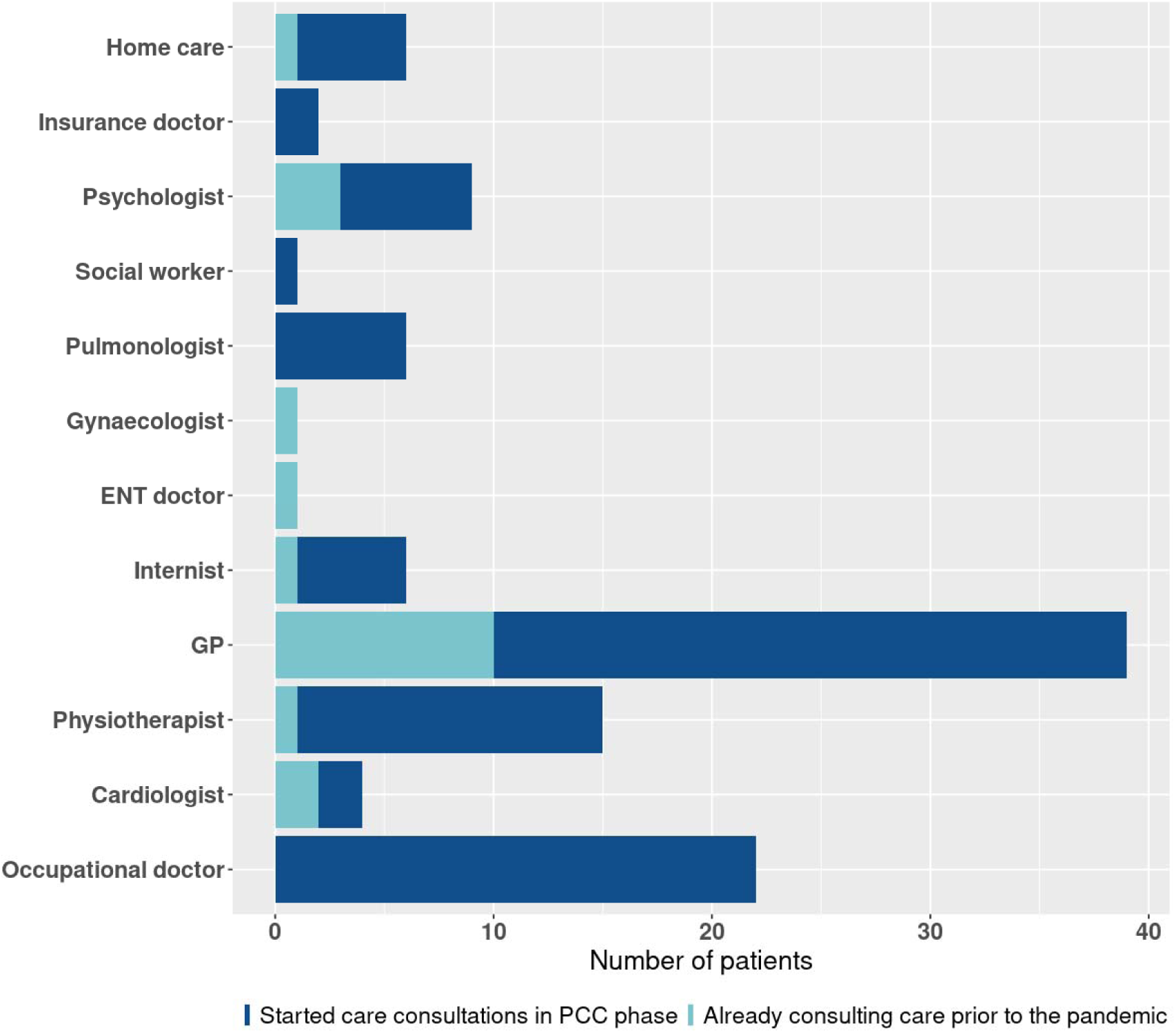
Care consultations during PCC for symptoms persisting after acute COVID-19, among 58 PCC patients on Bonaire, Caribbean Netherlands, who tested positive for SARS-CoV-2 between 1 March 2020 and 1 October 2021, by type of professional and prepandemic patient status.

Of patients consulting care during PCC, half (47%) were already under care prior to the pandemic (**Table 1)**, of whom the majority was using primary care **(Figure 1**). Considering the dynamics of care, our results show increased use of care provided by GPs (74% of patients consulting GP care during PCC were not in GP care prior to the pandemic) and home care (83% of patients were new users of home care), as well as care which requires a referral, such as the cardiologist (50% not under care prior to the pandemic), psychologist (67%), internist (87%), and physiotherapist (93%, respectively). One in three patients were consulting two professionals, and one in ten patients were consulting three professionals for their PCC symptoms. Excluding primary care, common combinations of care included the occupational doctor and physiotherapist; occupational doctor and pulmonologist; internist and pulmonologist; or psychologist and physiotherapist.

#### Factors correlated with consulting care during PCC

Patients living with other adults were over six times more likely to consult care during PCC compared to patients living alone (OR=6.11, 95% CI [1.30, 36.21]). There were no significant differences for other household compositions. Patients who were aware that their insurance covered certain medical costs for COVID-19 (after)care were four times more likely to consult care during PCC (OR=3.99, p<0.01, 95% CI [1.87, 8.81]). Consulting care during PCC was also significantly associated with comorbidity (OR=2.90, p<0.01, 95% CI [1.40-5.53]), pre-pandemic healthcare utilization (OR=2.52, p<0.01, 95% CI [1.30-4.95]), and having been hospitalized during the acute phase (OR=9.16, p<0.01, 95% CI [3.41, 29.21]) (**Table 1**).

#### Traditional healing and self-care practices

Roughly two out of three patients practiced self-care for persisting COVID-19 symptoms (**Table 1**). Ten themes were identified among patients’ descriptions of self-management practices for post-acute symptoms: Traditional healing practices, natural ingredients, use of supplements, healthy lifestyle, steaming, use of medication, following hygiene measures, memory aid, physical aid, and massage. Traditional healing practices were mentioned as one of the main ways of self-managing lingering symptoms. Local medicinal herbs (‘yerba di hole’ in Papiamentu), often from their own garden, were used in teas or other ways. Natural ingredients such as ginger, lemon, oregano, lemongrass, garlic, celery, onion, eucalyptus, curcuma, and honey were also used commonly.

Many patients described improving their lifestyle by making healthier nutritional choices, increasing physical activity, and getting more rest. Use of medication and supplements (vitamin C, zinc, and magnesium) appeared popular too. Medication used for treating symptoms was limited to over-the-counter medicines such as cough syrup, nasal spray, or painkillers (paracetamol, aspirin, diclofenac, codeine, naproxen).

> *“Yes, herbal tea, I drank smoothies, took extra vitamins A through Z, I rest more often, and have decided to listen to my body. In the past I used to ignore the signs, but now I step on the brakes.”*

> (Female, RTW before recovering).

#### Healthcare needs & barriers

Patients described a variety of needs related to COVID-19 (after-)care. Overall, patients expressed the need for more (non-pharmaceutical) attention by their doctor, shorter waiting times for specialist care, the need for a full check-up by their GP, lack of trust in medical results, or wishing their GP had not limited physical exams to the most severe cases. Second to improved medical care, most patients expressed the need for more information by and contact with the public health department. Several patients wished they were contacted more frequently, felt like they lacked information (specifically, information on testing policy, duration of COVID-19 symptoms, or vaccine effectiveness), and others mentioned missing emotional support or contact with a social worker to check how they were doing mentally:

> *“When you call your doctor, they only tell you to take medication and painkillers and tell you nothing else. I went to the doctor myself, I had a lot of pain and am very tired. When I come home from work I am often that tired that I only want to go to bed. I didn’t have these symptoms as badly before getting COVID-19. When I have a free day I can do nothing else, I only sleep. The GP now referred me to the internist and rheumatologist, but I can’t be helped until February. The rheumatologist is coming here in six months.”*

> (Female, RTW before recovering).

> *“They were afraid to examine the patient. They stayed at a distance a lot, which made the situation very impersonal. Only with severe symptoms you would be approached.”*

> (Female, RTW after recovering).

The physiotherapist was visited by a larger proportion of patients using care during the post-acute phase (21%) as compared to pre-pandemic (10%) (**Table 2**). Of patients who elaborated on their needs regarding physiotherapy and rehabilitative care, access to and duration of rehabilitative therapy formed barriers to their recovery. One patient explained it was unclear how to access rehabilitative programs, only learning about this option through their occupational doctor:

> *“I think the care is well organized. You just don’t know by yourself which paths to walk. I was pointed out by my occupational doctor that there is a treatment program at Bon Bida. After having been there once, I have already learned a lot to improve my lingering symptoms and to work on them.”*

> (Female, RTW before recovering).

### Return to work (RTW)

Of 129 PCC working age patients with pre-pandemic employment, 104 (81%) had RTW at the time of interview, of whom 68% had RTW within one month and 14% had RTW within two months (**Table 3**). One in three patients had RTW before feeling fully recovered, of whom 54% returned to the same working hours, 14% returning partly when not being fully recovered, and another 14% had started with reduced hours and built up to the same hours as prior to the pandemic while not being fully recovered. 77% of patients were not aware that their insurance covered certain COVID-19 related rehabilitative medical costs such as physiotherapy or a nutritionist.

**Table 3.**
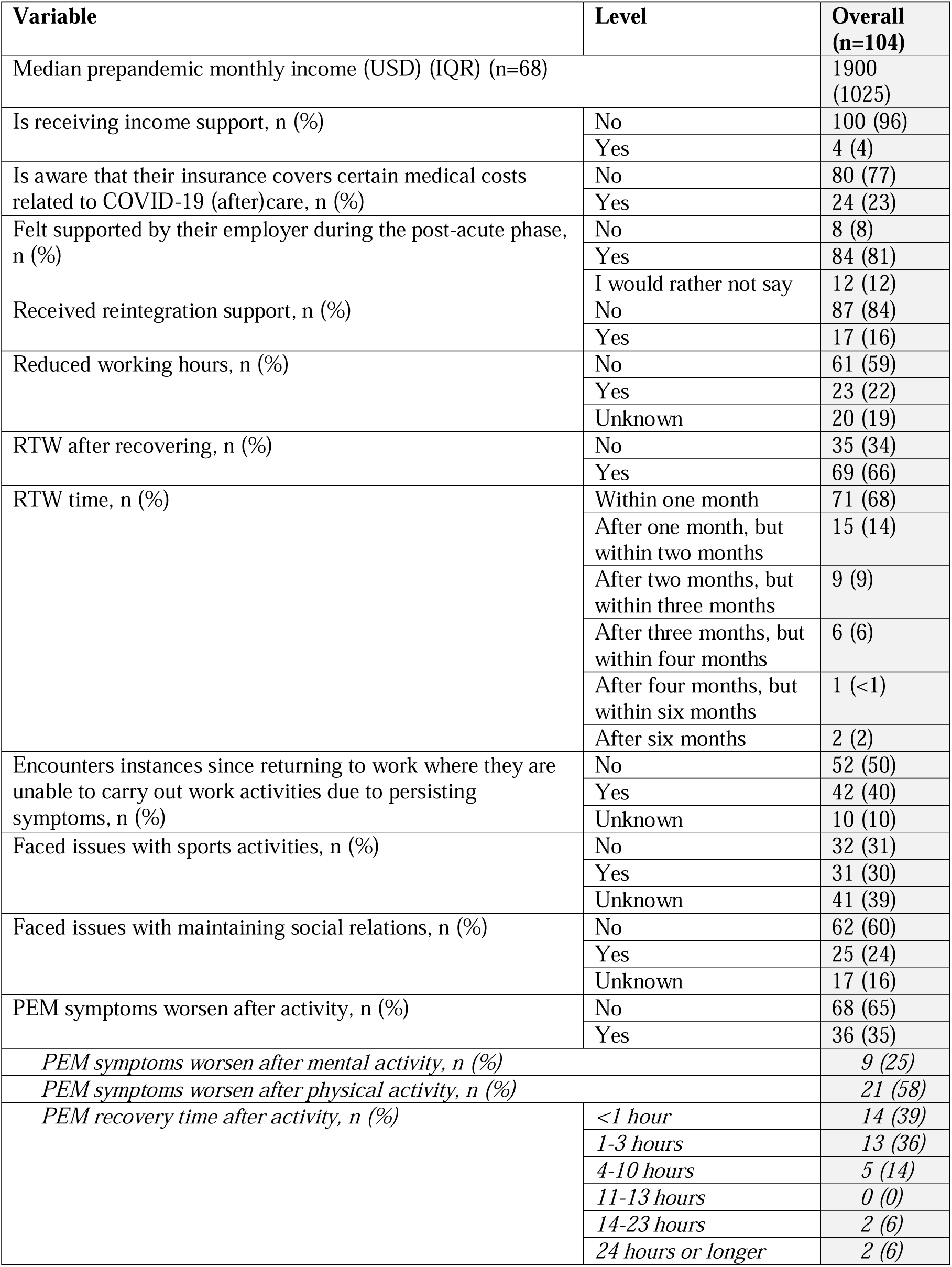
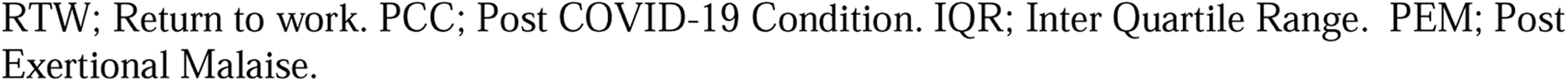
Daily functioning and return to work (RTW) conditions among a subgroup of 104 post COVID-19 condition (PCC) working age patients with pre-pandemic employment on Bonaire, Caribbean Netherlands, who tested positive for SARS-CoV-2 between 1 March 2020 and 1 October 2021 and who RTW at time of interview.

#### Activity limitations

In terms of performance at work, about one in five patients (22%) were working reduced hours and 40% of patients expressed that since having COVID-19, they were unable to carry out certain work activities. 35% of patients reported experiencing worsening of symptoms after any activity, of whom over half (58%) experienced worsening of symptoms after minimal physical activity and about one in four (24%) after minimal cognitive activity. About one in three patients (30%) experienced issues with carrying out sports activities since COVID-19 and one in four with maintaining social contacts (**Table 3**).

#### Occupational support

Most patients who RTW felt supported by their employer and/or colleagues when they were experiencing persisting symptoms (81%) (**Table 3**). 16% of patients received reintegration support, and a small group (4%) of patients were receiving income support. From open ended questions, we identified six themes related to support around reintegration at work: Support at home (sending fruit basket, delivering groceries), (lack of) compassion by colleagues/employer, financial (in)security, (lack of) contact by employer/colleagues, flexible return to work conditions, and medical help arranged by the employer.

Patients described a variety of occupational support systems beneficial to their recovery whilst on sick leave (mean age 42 years old). Most patients who felt supported recalled being contacted (frequently) by their employer and/or colleagues, receiving compassion, and being able to maintain financial security. Aside from these, some patients pointed out their employer provided flexible RTW conditions, allowing them to RTW with less hours, flexible working days, being allowed to work from home, or being allowed to start the working day later when experiencing worsened symptoms.

Overall, financial insecurity appeared an important theme amongst patients who did not feel supported. Some patients expressed they were never contacted or asked how they were feeling, whilst others felt misunderstood by their employer. Several participants suggested shortage of staff may have played a role in their employers’ flexibility in RTW.

> *“I received many messages and calls, and they did not stop paying my salary.”*

> (Male, RTW after recovering).

> *“When I was sick, colleagues did groceries for me and asked me how I was doing. I felt supported a bit by my employer, because they were very flexible with the days that I work. But they also wanted me to return to fulltime again, due to the shortage of teachers in the education sector.”*

> (Female, RTW before recovering).

## DISCUSSION

### Summary

Persisting physical and cognitive symptoms may very well limit patients’ ability to RTW or ability to concentrate on tasks at hand, having serious implications for job security, work productivity, and occupational safety and health [8]. Therefore, we examined RTW and care consultations during PCC by PCC patients on Bonaire, Caribbean Netherlands, and highlight patient identified needs related to support in the workplace and COVID-19 aftercare. Our findings show that, once diagnosed and referred, about one in three patients consulted care during PCC (37%). We found that not all patients with persisting symptoms were likely to seek care, and identified various barriers that patients experienced in their recovery trajectory. Secondly, we found a labor force participation reduction of 20%. Most patients opted to RTW shortly after having COVID-19, despite experiencing numerous limitations on daily activities, work performance, and ability to maintain social relations. Overall, our findings underline the relevance of viewing health issues within a broader socio-economic lens.

### Barriers

We explored whether financial and occupational support, health-related factors, and demographic factors played a role in patients care consultations during PCC and their activity limitations upon RTW. We found that patients who were aware that their insurance covered certain medical costs related to COVID-19 (after)care were nearly four times as likely to use care beyond the acute phase, as compared to patients unaware. In contrast, receiving income support was not significantly associated with consulting care during PCC.

Several patients expressed they missed guidance on how to navigate rehabilitative (PCC) care as well as general information on COVID-19 testing policies, duration of COVID-19 symptoms, or vaccine effectiveness in protecting against PCC. These findings suggest effective health communication may play a vital role in enabling patients to seek appropriate care, a need reiterated in <u>Dookeeram et al.</u> <u>(2023</u>), calling for policymakers in the Caribbean to better streamline access to information and resources to community members.

In our study of PCC prevalence and symptomatology on Bonaire [11], we found some PCC symptoms were more common among previously hospitalized PCC patients than among those that stayed at home, which may be one reason why care consultations during the PCC phase were higher among previously hospitalized patients. Another reason may be related to the COVID-19 related health insurance policies on Bonaire. Various studies have shown a relapsing-remitting pattern in the symptomatology and severity of PCC symptoms [9]. On Bonaire, patients whose condition worsened after experiencing a mild initial infection were ineligible for coverage of healthcare expenses [13], as only patients with a severe acute infection were eligible for COVID-19 aftercare. We believe this may be another explanation as to why we did not observe a significant uptake of care consultations during PCC by patients visiting their GP in the acute phase, but did find that patients who had been hospitalized during the acute phase were over nine times as likely to use care during PCC.

Additionally, our findings of care consultations during PCC being higher among hospitalized patients may also be explained by the referral policy in place in Bonaire’s hospital at the time: All patients admitted with/due to COVID-19 were scheduled for a follow-up with the treating specialist at four weeks after discharge and referred to a physiotherapist/nutritionist if there was an indication. For patients already under treatment by a physiotherapist/nutritionist during admission, treatment was continued after discharge.

Consulting care following the acute infection may have also been hampered by communication barriers between primary and secondary care on the island. During the study period, primary care physicians actively referred patients with persisting symptoms fitting the long-COVID case definition at the time, to the available COVID-19 aftercare on Bonaire. However, these physicians were not always (timely) informed of their patients’ history of COVID-19 related admission to the local hospital due to logistical issues within the communications infrastructure between primary and secondary healthcare providers; this, we believe, is likely to have hampered timely COVID-19 aftercare referrals through primary care.

### Healthcare use

We found that roughly one in ten PCC patients consulted a GP during the initial four weeks after infection. These patients often presented with fatigue (93%), cough (93%), and sleeping problems (86%). Secondly, we found that 37% of PCC patients in our study received care beyond the acute phase from at least one professional, of whom over half consulted a GP, one in three an occupational doctor, and roughly one in five sought treatment from a physiotherapist. These findings overlap with results from <u>Angarita-Fonseca et al. (2023</u>), who, among a Latin American study population, found that 33% of patients consulted the GP due to persisting symptoms [5]. Additionally, we found that traditional healing practices, such as the use of local herbs in teas, were common in self-managing persisting symptoms. These findings align with <u>Bardosh et al. (2023)</u>, who found the use of local teas and ingredients (such as ginger, garlic, and onions) was believed to have kept immune systems strong and severity low among individuals from Haiti throughout the 2020 lockdown [14].

Our findings show some patients seek a combination of care by different providers for treating their PCC symptoms. Patients living with other adults were over six times more likely to consult care during PCC compared to patients living alone (OR=6.11, 95% CI [1.30, 36.21]). We hypothesize adults living with other adults may have been more exposed to information and support regarding PCC, as compared to those living alone. When seeking PCC care, most patients expressed facing long waiting times for specialist care and found both psychosocial and medical support from GPs inadequate, describing physical exams were limited to those with severe symptoms. A small group of patients (5%) reported consulting a psychologist during PCC, where others described missing emotional support or contact with a social worker to check how they were doing mentally. We believe this low use of psychological care may be partially driven by mental health stigma rather than accurately reflecting PCC related mental healthcare needs among our study population, as much like on other Caribbean islands [6,15], mental health stigma on Bonaire remains a strong barrier to seeking care [16].

Overall, caution should be taken in interpreting our findings on care use during PCC as actual long-term care needs. We believe our findings present a limited capture of what the PCC burden on the Bonaire’s healthcare system may look like. As previously described, barriers such as restricted access to healthcare throughout waves of the pandemic, limitations in health insurance coverage as well as limited awareness of the existing cover, resumption of economic activity, availability of governmental subsidies for specific patient groups, or preference for self-managing symptoms by using home remedies [14] are likely to have influenced care consultations during PCC [3, 5, 17,18].

### Return to work

We found that workforce participation among PCC patients with pre-pandemic employment dropped by 19%. The majority of PCC patients RTW within one month (68%), and roughly one in five patients reduced their hours (22%), which was lower than found in other Latin American countries, where roughly one in three patients decreased time spent at work, school, and other activities [5].

Secondly, we found a large group of patients experienced activity limitations in some form whilst having RTW: 40% of patients who RTW experienced instances where they had problems in carrying out activities at work due to persisting COVID-19 symptoms, and 35% experienced worsening of their symptoms after carrying out any physical or cognitive activity. Outside of the workplace, about one in four patients faced issues in either sports activities or maintaining social relations since COVID-19.

There seem to be two sides to the coin of individual level economic impact [19] of (persisting) COVID-19: On one hand, patients experiencing functional limitations or not feeling fully recovered may be unable to afford to take sick leave when financial compensation is not regulated in work policies and such choices could lead to loss of income (leading to presenteeism). Despite the majority of these patients in our study feeling supported by their employer (81%), only a small group of patients received formal forms of support, such as reintegration support (16%) or income support (4%); and only one in four was aware that their insurance covered certain costs related to COVID-19 (after)care.

On the other hand, insurance coverage of COVID-19 (after)care on Bonaire has been limited to patients fulfilling specific criteria [13], which may also have driven patients to RTW prior to recovering in order to balance out additional care costs [5]. We found one in three patients RTW before feeling recovered from their symptoms and hypothesize that the economic burden of such policies may have driven some patients to RTW despite their symptoms, and to rely on other support systems in their environment. Caution should be therefore taken in interpreting our findings, which may falsely imply resilience [17] rather than patients opting not to take sick leave, as this may interfere with their job security and ability to generate income.

### Strengths & Limitations

To our knowledge, this is the first study to examine economic impact and activity limitations of PCC as well as related healthcare utilization among a (Dutch) Caribbean population. We were unable to examine income as a driver for consulting care during PCC or for RTW before recovering, as the many missings (n=70/157) in the income data left us with insufficient power to examine such correlations. We were unable to examine how long PCC patients continue to use care throughout the post-acute phase or what determines the moment of seeking care for persisting symptoms, as we did not include this in our survey. Additionally, we were unable to examine how long patients who were a candidate for hospital admission during the acute or post-acute phase, but chose to recover at home, were unable to RTW, as we did not include survey questions that would be required to identify this patient group. Regarding RTW, we did not include pre-pandemic working hours and could therefore not quantify the reduction in working hours. We did not correct for follow-up time in our RTW analyses. Follow-up time differed per patient, which may have impacted our RTW rates as these are partly determined by how much time patients have had to RTW since disease onset. However, as time since disease onset was quite long (see findings in our overarching prevalence paper [11]), we expect the impact of this to be minimal. Lastly, as we did not include occupation in our questionnaire, we were unable to consider if RTW rates could differ by sector.

### Implications for policy and future research

Existing health disparities, combined with increasing rates of non-communicable diseases and constrained health systems have fueled the COVID-19 pandemic in the Caribbean region [4]. Although the association between racial differences and COVID-19 outcomes have been well established in other LMIC settings [20], research of how PCC has affected disparities in the Caribbean is lagging behind [21]. Few European Dutch studies of COVID-19 or PCC have thus far included non-white racial groups in their study population, and the Dutch overseas territories are often excluded from European Dutch public health laws, policies, and research opportunities due to current political structures [18]. This is a major constraint for public health within the Kingdom, as results of European Dutch research can rarely be generalized to the Dutch Caribbean settings due to the study population not being representative. We believe this makes our findings of essential value to the region to gain insights into the longer term impacts of COVID-19 and emphasizes the need to continue including Dutch Caribbean populations into future research on PCC care and support strategies as well as broader pandemic preparedness plans stemming from COVID-19 impact evaluations in the Region.

Our study includes data of persons who received a laboratory confirmed SARS-CoV-2 positive test result up until 1 October 2021. Since then, SARS-CoV-2 has continued to circulate on Bonaire [22] and therefore PCC and the accompanying burden on Bonaire’s healthcare system, as well as the economic burden, has likely risen in the period immediately after our study was conducted. Our findings add argument to promoting a multidisciplinary approach to COVID-19 aftercare, since PCC patients can enter the healthcare system via different healthcare professionals [5]. We believe local health professionals may play a vital role in advocating for policy changes related to COVID-19 aftercare, based on their real-life experiences with this patient population throughout the pandemic. Additionally, we believe including various social and economic aspects related to recovery from COVID-19 highlighted in our study into existing PCC care on Bonaire will benefit patients’ recovery trajectory.

### Conclusions

Our findings shed light on the labor force implications and healthcare burden of PCC, suggesting labor force participation of working age PCC patients dropped by 20% on Bonaire, Caribbean Netherlands. Additionally, we highlight various factors related to care use during PCC by patients on Bonaire and patient identified needs related to support in the workplace and COVID-19 aftercare. Improving occupational and healthcare policies and communication between healthcare professionals and PCC patients on Bonaire may provide various benefits for recovering patients.

## Acknowledgements

We would like to thank Alicia Krijgsman and Karin Cox (Public Health Department Bonaire), members of CBS Bonaire, and interviewers of the Tempo team on Bonaire for their contributions in the design and preparation of the overarching study, data collection, and logistical assistance with implementing the overarching study on Bonaire. We would like to thank Caroline van den Ende (RIVM, Cib, EPI) for advising on the latest publications regarding PCC and Thomas Dalhuisen (RIVM, Cib, EPI) for assistance in data management for the overarching study.

## Authors contributions

DSF Berry: Conceptualization, data curation, methodology, software, resources, formal analysis, thematic analysis, visualization, writing – original draft; submitting author.

G Marchena: Conceptualization, thematic analysis, reviewing – original draft. A Veggh: Conceptualization, reviewing – original draft.

I Tiemessen: Investigation, data curation.

L Jaspers: Funding acquisition, data curation, project administration, reviewing – original draft.

E Geubbels: Conceptualization, methodology, supervision, visualization, writing – original draft.

## Data Availability Statement

Underlying dataset and code are available upon request.

---

**Supplementary Table 1.** Description of variables included in the analyses.

| Variable | Type | Description |
| --- | --- | --- |
| Age | Continuous | Age in years. |
| Gender | Binary | Male, female. |
| Education level | Categorical | Categories are: Low level education (no education, primary level education, lower vocational & lower secondary education); moderate level education ( higher secondary education); high level education (university level & post-secondary education). |
| Presence of one or more comorbidities | Binary | Binary variable indicating whether a person suffers from any of the following prior to the start of the pandemic: self-reported post-acute respiratory problems (including asthma, chronic obstructive pulmonary disorder, tuberculosis), post-acute cardiovascular disease, diabetes, severe kidney disease (including dialysis or kidney transplantation), HIV, severe liver disease, being immunocompromised, cancer, depression or anxiety, stroke, rheumatism, arthritis, or memory loss due to a neurological condition or dementia. |
| Number of symptoms in the acute phase | Numerical | Number of symptoms a patient experienced during the acute phase of their SARS-CoV-2 infection. Symptoms include: chest pain, concentration problems, cough, fatigue, headache, heart palpitations, loss of appetite, reduced muscle strength, loss of sense of smell, loss of sense of taste, muscle ache, reduced physical endurance, shortness of breath, sleeping problems. |
| Monthly income | Continuous | Individual monthly income after taxes, in US Dollars. Income prior to the start of the pandemic. |
| Employment status | Categorical | Categorical variable indicating whether a person was employed, student, or unemployed prior to the pandemic. |
| Household composition | Categorical | Categories are: Living alone; living with partner and kids; living with other adults (e.g. friends, roommates), living with parent(s); living with partner no kids; multigenerational household; nursing home resident; single parent. |
| COVID-19 symptoms | Binary | Binary variables indicating whether a person experienced each of the following symptoms during the acute/post-acute phase: Shortness of breath, cough, worsened physical endurance, chest pain, loss of muscle strength, muscle ache, loss of sense of smell, loss of sense of taste, loss of appetite, heart palpitations, concentration problems, sleeping problems, fatigue, headache. |
| GP visit in the acute phase | Binary | Binary variable indicating whether a patient visited or was visited by a GP during the acute phase of their SARS-CoV-2 infection. |
| Hospitalized during the acute phase | Binary | Binary variable indicating whether a patient was hospitalized during the acute phase of their SARS-CoV-2 infection. |
| Prepandemic healthcare utilization | Binary | Binary variable indicating whether a patient visited a (para)medical healthcare professional prior to the start of the pandemic. |
| Prepandemic medical healthcare utilization | Categorical | Categorical variable indicating whether a patient visited a one of the following medical healthcare professionals prior to the start of the pandemic: General practitioner (GP), occupational doctor, pulmonologist, cardiologist, ENT doctor, internist, surgeon, gastroenterologist, nephrologist, dermatologist, geriatric doctor, neurologist, eye doctor, orthopedist, rheumatologist, urologist, gynecologist. |
| Prepandemic paramedical healthcare utilization | Categorical | Categorical variable indicating whether a patient visited a one of the following paramedical healthcare professionals prior to the start of the pandemic: Physiotherapist, manual therapist, osteopath, occupational therapist, psychologist, mental health worker, social worker, home care. |
| PEM symptoms after any activity | Binary | Binary variable indicating whether a person experienced worsening of long-term symptoms after any activity. |
| PEM symptoms after physical activity | Binary | Binary variable indicating whether a person experienced worsening of long-term symptoms after minimal physical activity/exercise, among patients who indicated they experienced worsening of symptoms after any activity. |
| PEM symptoms after cognitive activity | Binary | Binary variable indicating whether a person experienced worsening of long-term symptoms after minimal cognitive activity/exercise, among patients who indicated they experienced worsening of symptoms after any activity. |
| PEM recovery time after activity | Categorical | Categorical variable indicating how long it took for symptoms to clear after experiencing worsening of long-term symptoms after minimal cognitive activity/exercise, among patients who indicated they experienced this: <1 hour, 1-3 hours, 4-10 hours, 11-13 hours, 14-23 hours, 24 hours or longer. |
| Mobility issues | Binary | Binary variable indicating whether a person ability to move around physically without mobility aids has been impacted as a consequence of experiencing COVID-19. |
| Problems at work | Binary | Binary variable indicating whether a person suffered from long-term problems in carrying out activities at work as a consequence of experiencing COVID-19. |
| Problems with sports | Binary | Binary variable indicating whether a person suffered from long-term problems in carrying out sports activities as a consequence of experiencing COVID-19. |
| Problems with social contacts | Binary | Binary variable indicating whether a person suffered from long-term problems in maintaining social contacts as a consequence of experiencing COVID-19. |
| Role at work | Binary | Binary variable indicating whether a person was (un)able to return to their role at work since experiencing long-term COVID-19 symptoms. |
| Reduced work hours | Binary | Binary variable indicating whether a person returned to work in reduced hours since experiencing long-term COVID-19 symptoms. |
| Return to work | Categorical | Categorical variable indicating after how much time a person returned to work since experiencing COVID-19: Within one month; within one month but after two months; within two months but after three months; within three months but after four months; within four months but after six months; after six months; have not (yet) returned to work. |
| Income support | Binary | Binary variable indicating whether a person is receiving government issued income support at the time of interview. This income support was issued by the government through the Dutch Ministry of Social Affairs and Employment (SZW) or by the Public Entity of Bonaire (OLB). |
| Reintegration support | Binary | Binary variable indicating whether a person received reintegration support from their employer when returning to work after having COVID-19. |
| Support work | Binary | Binary variable indicating whether a person felt supported by their employer and work environment when experiencing long-term symptoms. |
| Healthcare use beyond the acute phase | Binary | Binary variable indicating whether a patient visited a (para)medical healthcare professional during the post-acute phase. Professionals included: Acupuncturist, occupational doctor, cardiologist, Cesar therapist, nutritionist, occupational therapist, physiotherapist, GP, homeopathic therapist, internist, ENT doctor, speech therapist, pulmonologist, social worker, manual therapist, psychologist/ psychotherapist, rehabilitation doctor, insurance doctor, home care. |
| Self-care | Binary | Binary variable indicating whether a person performed any self-care activities when experiencing post-acute symptoms. |
| Insurance | Binary | Binary variable indicating whether a person was aware of the insurance policy related to coverage of certain rehabilitative care for COVID-19 patients, including physiotherapy and nutritionist, through the insurance provider. |
| Vaccination status prior to SARS-CoV-2 infection | Binary | Binary variable indicating whether a person was vaccinated with at least one dose of the Pfizer/BioNTech/Comirnaty vaccine at least 14 days prior to their SARS-CoV-2 infection, which was the only vaccine used for COVID-19 vaccinations on Bonaire during the inclusion period of this study. |
| Recovery status | Binary | Binary variable indicating whether a person had recovered at the time of interview. |

PEM; Post Exertional Malaise.

**Supplementary Table 2.**
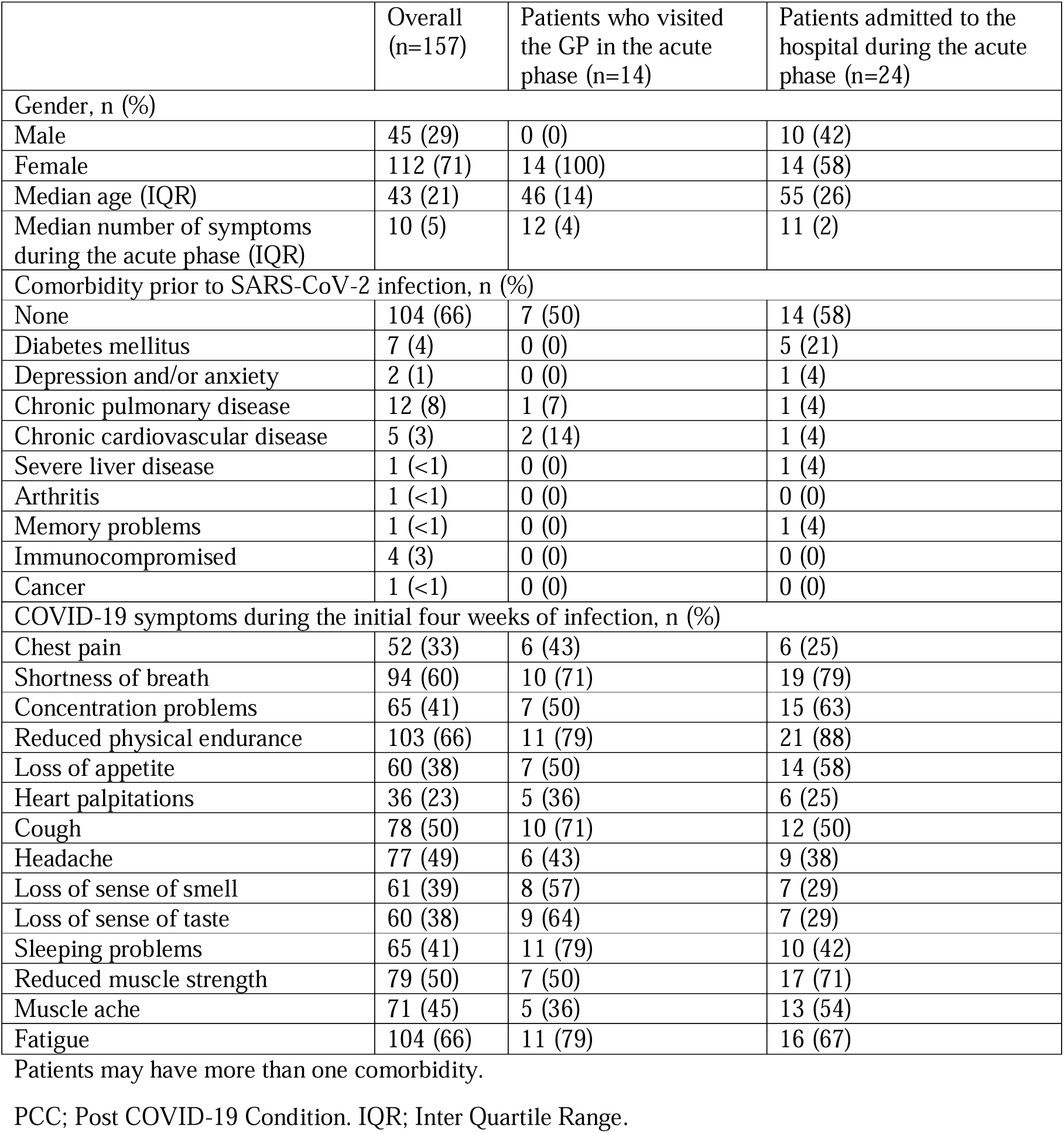
Characteristics of PCC patients on Bonaire, Caribbean Netherlands, who used care during the initial four weeks after COVID-19 disease onset.

